# Evaluation of ID NOW and RT-PCR for Detection of SARS-CoV-2 in an Ambulatory Population

**DOI:** 10.1101/2020.12.07.20245225

**Authors:** Yuan-Po Tu, Jameel Iqbal, Timothy O’Leary

## Abstract

Diagnosis of the SARS-CoV-2 (COVID-19) requires confirmation by Reverse-Transcription Polymerase Chain Reaction (RT-PCR). Abbott ID NOW provides fast results but has been criticized for low sensitivity. Here we determine the sensitivity of ID NOW in an ambulatory population presenting for testing. The study enrolled 785 symptomatic patients, 21 of whom were positive by both ID NOW and RT-PCR, and 2 only by RT-PCR. All 189 asymptomatic patients tested negative. The positive percent agreement between the ID NOW assay and the RT-PCR assay was 91.3%, and negative percent agreement was 100%. The results from the current study were included into a larger systematic review of literature where at least 20 subjects were simultaneously tested using ID NOW and RT-PCR. The overall sensitivity for ID NOW assay was calculated at 84% (95% CI 55-96%), and had the highest correlation to RT-PCR at viral loads most likely to be associated with transmissible infections.

## Introduction

The SARS-CoV-2 (COVID-19) virus has infected over sixty-three million persons worldwide, causing over 1, 500,000 deaths as of December 1, 2020. Infected individuals may be asymptomatic, or may have a range of symptoms varying from a mild upper respiratory illness or gastrointestinal distress to severe respiratory distress with multisystem failure and death^1^. Definitive diagnosis requires laboratory detection of virus, and is required for patients to be eligible for both clinical trials and current antiviral drugs and biologicals approved by the Food and Drug Administration (FDA) under Emergency Use Authorization (EUA)^2^. Early in the pandemic, detection of SARS-CoV-2 relied predominately on Reverse Transcriptase Polymerase Chain Reaction (RT-PCR) assays performed in moderate to high complexity CLIA-certified laboratories. RT-PCR assays performed in certified laboratories are highly sensitive and specific but require expensive and complex analyzers operated by certified and highly skilled laboratory workers; in many cases, these tests have required turnaround times of nearly a week or more.

The use of testing strategies with a rapid turnaround may allow for an earlier detection and better isolation of confirmed cases compared to laboratory-based diagnostic methods, as well as facilitate earlier treatment decisions and provide guidance on appropriate use of personal protective equipment. On March 27, 2020, Emergency Use Authorization was granted for the COVID-19 EUA assay on the ID NOW system (Abbott, Scarborough Diagnostics). The ID NOW system is a point-of-care (POC) device that uses an isothermal nucleic acid amplification technique to allow for nucleic acid amplification without thermal cyclers and allows for results to be obtained quickly. The ID NOW SARS-CoV-2 assay (Abbott) amplifies a unique region of the RdRp genome with a manufacturer’s claimed limit of detection (LOD) of 125 genome equivalents/mL. The isothermal technique allows for positive results to be available as soon as 5 minutes into the assay, and negative results within 13 minutes.

Since its release, several studies have been published demonstrating a sensitivity relative to RT-PCR from 44% to 94% percent (excluding a study with only a single positive). Studies have shown fairly definitively that the limit of detection for ID NOW COVID-19 requires significantly higher amounts of virus than most RT-PCR assays^3,4^, but the clinical importance of this finding has been tempered by the observation that virus detectable only at high cycle time threshold (Ct) values is generally not culturable, and may therefore not be sufficiently high to infect others^5^. Additional studies have suggested that nasal viral loads peak at around the time symptoms appear, and fall off as infection lingers^6^. Hence, a diagnostic approach that is adequate early in the course of infection may be inadequate for patients that present later in the course of disease. Thus, the decision on whether the time-advantage of a lower sensitivity device offsets the potentially higher limit of detection may depend on the context in which that device is employed.

To better understand the performance characteristics and tradeoffs involved in the use of the ID NOW system, we have carried out a prospective clinical evaluation of the ID NOW system in the context of a community screening program focusing on symptomatic persons demonstrating one or more clinical feature of SARS-CoV-2 infection, comparing the results with those obtained by RT-PCR testing. We have augmented the findings of this investigation with a systematic review and meta-analysis of ID NOW performance, focusing on ambulatory community populations undergoing initial testing.

## Methods

### Study Population and Sample Collection

#### Clinical Study

The IRB-approved clinical study was conducted at The Everett Clinic between April 8^th^ and 22^nd^, 2020, and engaged ambulatory symptomatic patients seen in the Febrile Upper Respiratory Infection (F/URI) clinics and other patients from non-F/URI clinics. Patients who were unable to demonstrate understanding of the study, not willing to commit to having all samples collected, had a history of nosebleed in the past 24 hours, nasal surgery in the past two weeks, chemotherapy treatment with documented low platelet and low white blood cell counts, or acute facial trauma were excluded; nonetheless, an attempt was made to consecutively enroll all eligible patients.

The original study design called for enrolling 2000 symptomatic and 500 asymptomatic subjects, which would have provided a power of 95% for finding a difference of 5% in the sensitivity of ID NOW compared with a reference standard. The study design assumed a population prevalence of 10%, and the study was terminated early when the population prevalence dropped to such a low level as to make the study unaffordable.

Patients who consented to the study had two sterile foam swabs (Puritan, #PK002196) obtained by trained clinical staff. To ensure maximum loading of viral material, each swab sampled in both anterior nares (AN). To ensure that both swabs had equal opportunity to collect viral material, (Figure 1) the collection of the two swabs used a cross-over method. The procedure is as follows:

1. The first swab was gently inserted into the right nostril until resistance was met at the level of the turbinate (less than one inch into the nostril) and gentle pressure was applied to the outside nasal wall and the swab was rotated several times against the nasal wall and then slowly remove from the nostril.
2. The second swab was gently inserted into the left nostril and sampling was obtain in a similar manner.
3. Next, the first swab was inserted into the left nostril and sampling was obtained in a similar manner.
4. Finally, the second swab was inserted into the right nostril and sampling was obtained in a similar manner.

**Figure 1:**
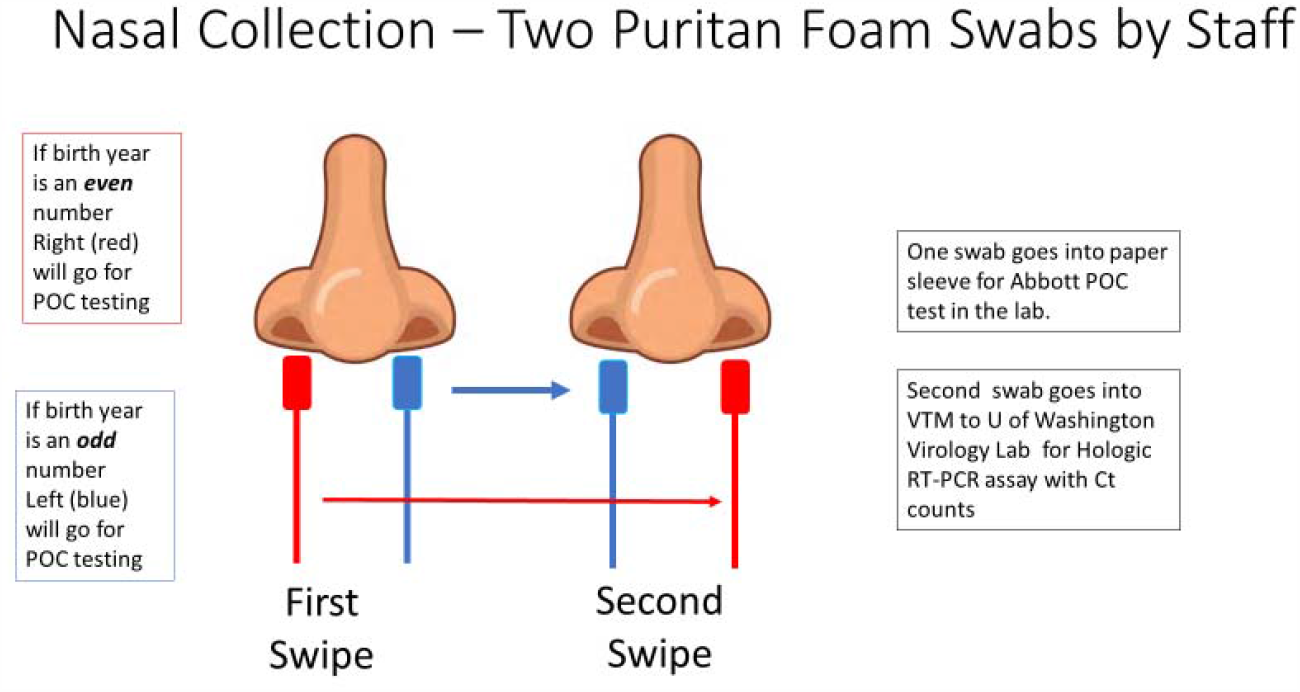
The collection methodology to ensure proper randomized sampling of nares for simultaneous analysis for SARS-CoV-2 by the ID NOW isothermal amplification and Hologic Panther RT-PCR assays. A total of two swabs was collected on each patient, with patients having an even birth year number the right nares was collected first followed by a second swipe in the left nares and then for ID NOW point-of-care (POC) testing (depicted as red swab). For those patients the other swab (blue swab) was sent for SARS-CoV-2 RT-PCR analysis by the Hologic Panther assay. For patients having an even birth year, the swabs sent for testing was reversed with the blue swab sent for ID NOW testing and the red swab sent for RT-PCR analysis.

If the patient’s year of birth ended in an even year, the first swab inserted into the right nostril was designated for SARS-CoV-2 testing using the point-of-care (POC) analyzer. If the patient’s year of birth was an odd year, the first swab inserted into the left nostril was designated for SARS-CoV-2 testing using the ID NOW analyzer. The swab designated for testing in the ID NOW analyzer was reinserted into the original paper sleeve packaging, a patient label was affixed, placed in a plastic bag and transported to the clinic lab on site for immediate testing. Typically, fewer than fifteen minutes passed between the time the room-temperature sample was collected and the time that the swab was inserted into the ID NOW sample receiver.

The remaining swab was placed in VTM (Medical Diagnostic Laboratories, L.L.C.). After a patient label was affixed the specimen was placed in a plastic bag and transported at 4 degrees centigrade to the University of Washington Virology Lab, where a Hologic Panther Fusion® SARS-CoV-2 assay (Marlborough, MA) was performed per manufacturer’s recommendations. With the Hologic assay, a sample is considered positive if an amplification signal is detected at a cycle time (Ct) of 42 cycles or less. Those involved in the RT-PCR assay were blinded to the ID NOW result.

ID NOW results which were reported as “invalid” were treated as negative when calculating the sensitivity of the ID NOW test; moreover, they were excluded from computations of specificity since this result would be expected to trigger reflex testing. Confidence intervals for sensitivity and specificity were calculated using Newcombe’s efficient score method^7^ (with continuity correction) as implemented in the Vassarstats calculator for confidence intervals of a proportion (http://vassarstats.net/).

#### Review and Meta-Analysis

Our systematic review was designed to answer two questions:

What is the limit of detection for the ID NOW assay?

What is the clinical sensitivity of the ID NOW SARS-CoV-2 assay in comparison with an RT-PCR assays for SARS-CoV-2?

The study is based on a protocol registered on PROSPERO (CRD42020204441), but a complete protocol has not been published. PubMed, medRxiv and bioRxiv were searched over the interval from January 1, 2020 to August 16^th^, 2020 using the search “ID NOW”, “isothermal amplification,” lamp isothermal”. Following the initial identification of papers, the titles and abstracts were screened to eliminate papers not meeting the prespecified inclusion criteria as defined below and diagrammed in Supplemental Figure 1. Papers remaining after this process were rescreened, particularly since many of the papers reviewed were in the form of research letters that did not have an abstract. Ultimately, 14 papers that met inclusion criteria for clinical comparison were available for analysis, as shown in the PRISMA flow diagram (Supplemental Figure 1). In addition, one additional paper addressing the limit of detection for ID NOW was identified.^8^

To be included in the systematic review, studies were required to include a minimum of 20 unique subjects. Studies must have compared samples obtained simultaneously from the same site, or from an equivalent site. Both split-sample designs and independent sample designs were considered. Results must have been reported in a manner that allowed construction a confusion matrix including the RT-PCR and ID NOW test. Because “discrepant analysis” provides biased sensitivity estimates, studies using this technique to resolve diagnostic conflicts between two sites were not to be included unless data could be analyzed independently of the discrepant analysis. If multiple time points were included in one of the included studies, only the first time point was to be used in our analysis. If confusion matrices could only be constructed from data involving multiple time points from the same patients, the study was excluded. No attempt was made to obtain data from the investigators involved in these published studies.

Study information was recorded on a predetermined data extraction form that included study author, type of study, inclusion and exclusion criteria, setting, sample types, swab types, transport medium, manufacturer or description of nucleic acid amplification assays, as well as space to record study results in the form of confusion matrices. The potential for bias associated with each study was evaluated using the QUADAS2 instrument. The risk of spectrum bias, which is the variability of medical test performance that happens when tests are given to different mixes of patients at different locations, was assessed from the perspective of testing as an initial diagnostic method; the risk estimate does not constitute a judgement on the quality of the study, which may have been performed to demonstrate assay validity, assessment of recovery, or other purposes different than that for which we evaluated potential bias.

Because the choice of any particular diagnostic device as a “gold standard” provides a biased estimate of relative sensitivity which compared with all other devices, a composite reference standard (CRS) was computed for each study on the basis of all devices and sample types included in the study, when possible. Equivocal results and assay failures were not used in the calculation of sensitivity, nor in the construction of the CRS for each study. Where multiple RT-PCR assays were performed, only the performance of the most sensitive of these assays (as measured using the composite reference standard) is reported in results tables. Confidence limits for sensitivity were computed using Newcombe’s efficient score method, as above. Criteria for performing a formal meta-analysis were prespecified as follows: 1) studies used the same amplification technology [such as RT-PCR] as a reference; 2) studies used the same upper airway sample site [AN, mid-turbinate (MT) and nasopharynx (NP)] could be included together, but not admixed with studies based on oropharynx samples); 3) studies enrolled a similar patient mix (e.g. symptomatic, asymptomatic, hospitalized) and similar clinical environment (drive-through/ community health center or hospital). Three papers in which with a low risk of bias were deemed appropriate to include in a meta-analysis were analyzed using a diagnostic effects model (der Simion – Laird) as implemented by OpenMetaAnalyst software program. The choice of any particular diagnostic device as a “gold standard” provides a biased estimate of relative sensitivity which compared with all other devices^9^. When two devices, each of which is expected to have a near-zero false positive rate, are being compared, the use of a composite reference standard (CRS) is a reasonable approach by which to reduce this bias^10^. For this reason, we compared the performance of ID NOW and RTPCR methods with a composite reference standard in which the specificity of all assays was considered to be perfect, and a positive result for any assay was considered to be a “true positive.” Equivocal results and assay failures were not used in the calculation of sensitivity, nor in the construction of the CRS for each study. Where multiple RTPCR assays were performed, only the performance of the most sensitive of these assays (as measured using the composite reference standard) is reported in results tables. Confidence limits for sensitivity were computed using Newcombe’s efficient score method, as above. Criteria for performing a formal meta-analysis were prespecified as follows: 1) studies used the same amplification technology [such as RT-PCR] as a reference; 2) studies used the same upper airway sample site (AN, MT and NP could be included together, but not admixed with studies based on OP samples; 3) studies enrolled a similar patient mix (e.g. symptomatic, asymptomatic, hospitalized) and similar clinical environment (drive-through/ community health center or hospital). Three papers in which with a low risk of bias were deemed appropriate to include in a meta-analysis were analyzed using a diagnostic effects model (der Simionian – Laird^11^) as implemented by OpenMetaAnalyst software program^12^. Since our model is built on the assumption that there are no false positive ID NOW results, a value of 0.5 was added to all cells as a continuity correction.

## Results

### Clinical Evaluation

The evaluation enrolled 785 symptomatic patients, 21 of whom tested positive for SARS-CoV-2 by both the ID NOW and Hologic assays, and 2 of whom tested positive only with the Hologic assay (Table 1). In addition, the evaluation enrolled 189 asymptomatic patients, none of whom tested positive by either ID NOW or RT-PCR. An “invalid” ID NOW assay result was reported for 9 subjects (2 asymptomatic, 7 symptomatic), all of whom tested negative by RT-PCR. Thus, the positive percent agreement between the ID NOW assay and the Hologic Panther Assay was 91.3%, and the negative percent agreement was 100%. The median cycle time (Ct) values in patients who had a positive Hologic RT-PCR was 28.2.

**Table 1:**
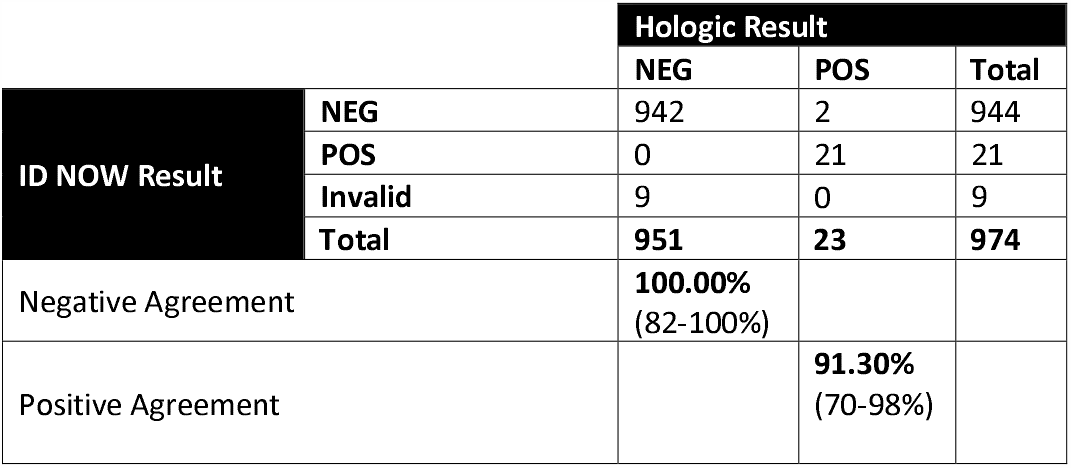
Results from the clinical evaluation comparing randomized anterior nares samples for the ID NOW compared to the Hologic Panther SARS-CoV-2 RT-PCR assay.

Two patients had discordant results with a negative ID NOW test and a positive Hologic RT-PCR test. The Hologic Ct values on the two discordant patients were 36.5 and 38.1. Of these discordant results, one patient is a 58-year-old woman who was a former smoker who presented with a cough and mild respiratory symptoms for approximately six weeks. She was retested four days after the initial discordant results at which time she tested negative in both the ID NOW and Hologic RT-PCR assays. The other patient with discordant results was a 34-year-old man with diabetes; he declined repeat testing but clinically was improving when contacted by phone.

### Systematic Review and Meta-Analysis

Forty papers were considered for inclusion. Of these, fourteen met inclusion criteria, as reflected in the PRISMA diagram (Supplemental Figure 1); nine of those 14 studies enrolled 100 or more subjects. A brief summary of the studies included in our review, including the clinical study reported in this paper, is described in Table 2. A brief discussion of each paper including the results used in this review is presented in the Supplemental materials.

**Table 2:**
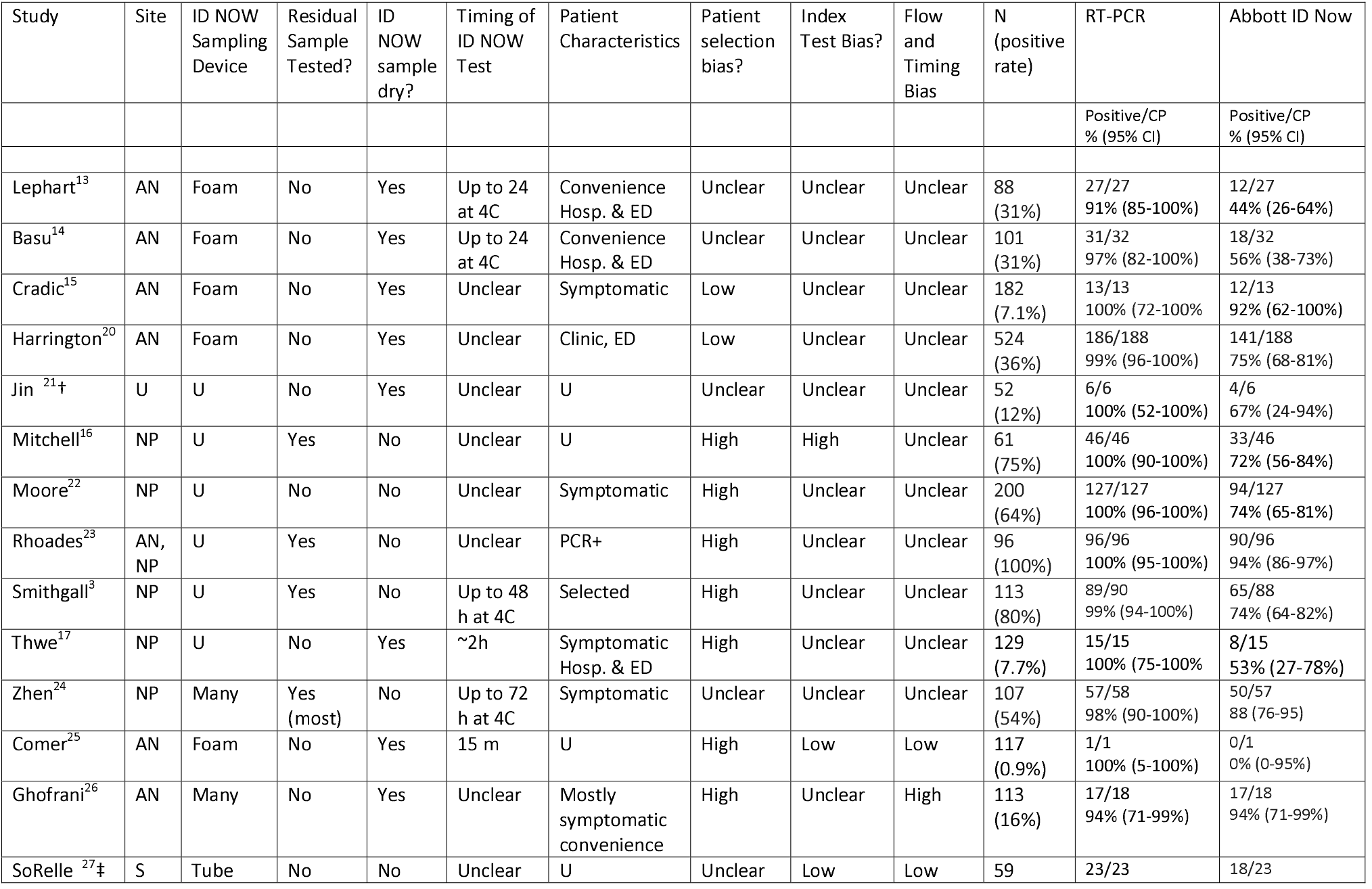
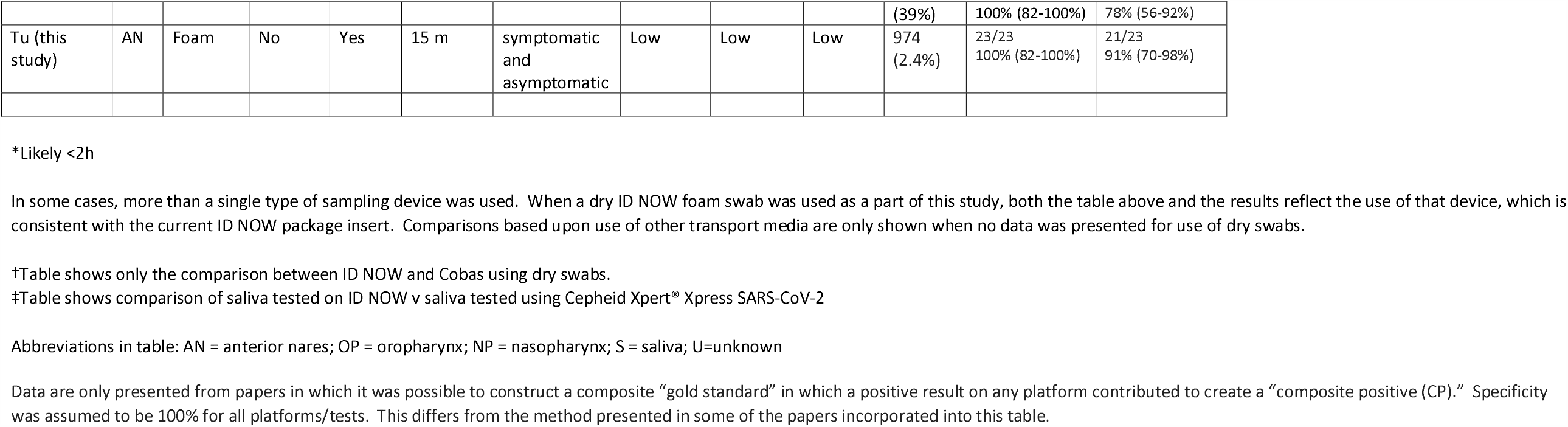
Studies Included in the Systematic Review

The risk of patient selection spectrum bias associated with the study population, or method of recruitment, was rated as either “high” or “unclear” for 12 of the published studies; this was the most common concern raised in the quality assessment. Studies with a high or unclear risk of bias were characterized by failure to present patient symptom status (five studies), inclusion of subjects who had previously tested positive for SARS-CoV-2 (one study) or use of investigator-selected or non-clinical convenience samples. Evidence of bias associated with the conduct of RT-PCR testing was not identified for any of the fourteen studies meeting inclusion criteria. Several studies suffered either from unclear or elevated risk of index test or flow and timing biases (detailed further in Supplementary Materials).

The clinical sensitivity of the ID NOW assay was lower than that of the RT-PCR assay, when both were compared to the composite reference standard, in 14 of the 15 studies shown in Table 2. In studies reporting more than a single positive RT-PCR result, the sensitivity of ID NOW, as compared to the composite reference standard, varied from 44-94%, while that of the RT-PCR test varied from 91-100%. For studies in which patient selection bias was rated low, the sensitivity of ID NOW (in comparison with the composite reference standard), ranged from 60-92% (Table 2). This corresponds with published analytical sensitivity estimates that have shown limits of detection for ID NOW that are several orders of magnitude higher than those of RT-PCR assays, ranging from 3900^13^ to 20,000^4^ gene copies/mL, and data published on an FDA web site (https://www.fda.gov/medical-devices/coronavirus-covid-19-and-medical-devices/sars-cov-2-reference-panel-comparative-data) that suggests a 500-fold higher limit of detection for the ID NOW platform than for the Panther Fusion Assay employed in our clinical study. These results are consistent with the studies in our systematic review that showed discordance among assays to be most frequent when Ct values were relatively high (see Supplemental Materials).^3,4,13–16^

The ID NOW instructions for use (IFU, https://www.fda.gov/media/136525/download) have changed over time, but generally have called for samples to be tested no later than one hour after specimen acquisition and kept at room temperature during that period. The changes in the IFU have made it difficult to assess if published studies provided sufficient information to allow a determination that conformation to instructions for use was followed sufficiently. Four studies included in this review were based upon a split/residual sample design; the calculated sensitivity for the ID NOW in these studies ranged from 72 to 94%. For eight of the studies, timing of the ID NOW test was unclear, while for 4 studies samples were held after collection at 4°C for up to 24 hours (2 studies), 48 hours (1 study), or 72 hours (1 study). The degree to which this affects assay sensitivity is unclear; however, it is noteworthly that a study that held samples for up to 72 hours reported ID NOW sensitivity (as compared to the composite reference standard) of 88%, while another study that held samples for no more than two hours reported a sensitivity of 56%. Only one of the studies captured for this systematic review reported a time-to-test for ID NOW of ≤1 hour, and that study included only one patient that tested positive using either device. Thus, there is no conclusive evidence that the refrigeration serves as an explanation for varying sensitivies.

There was no obvious relationship between the sample site, such as anterior nares (AN) versus nasopharynx (NP), or sampling device and the sensitivity of the ID NOW test. Both high and low concordance with the composite reference standard were found for both sites and for both foam and flocked swabs. Similarly, both good performance and poor performance was found for both samples transported in a medium, or transported dry. Finally, the overall prevalence of positive findings in the study population was not correlated with the performance of ID NOW in the studies we have examined.

We included the two cohorts with low risk of patient selection bias, together with the current study, in a meta-analysis, the results of which are shown as forest plots in Figure 2. The sensitivity of ID NOW, as compared with the reference standard, was estimated at 82% (Figure 2A); the lower and upper 95% confidence bounds were 67% and 91%, respectively. Measures of heterogeneity did not reach statistical significance (τ^2^ =0.25, Q[df=2] = 3.67, p=0.16, I^2^=45.53). In contrast the sensitivity of RT-PCR (Figure 2B) was estimated at 98% with a 95% CI of 96-99%. There was no suggestion of heterogeneity (τ^2^ =0.000, Q[df=2] = 0.453, p=0.112, I^2^=0.000). The sensitivity estimates for both ID NOW and RT-PCR were reduced, probably by about 2%, by the need to include a continuity correction in the der Simionian – Laird computations.

**Figure 2:**
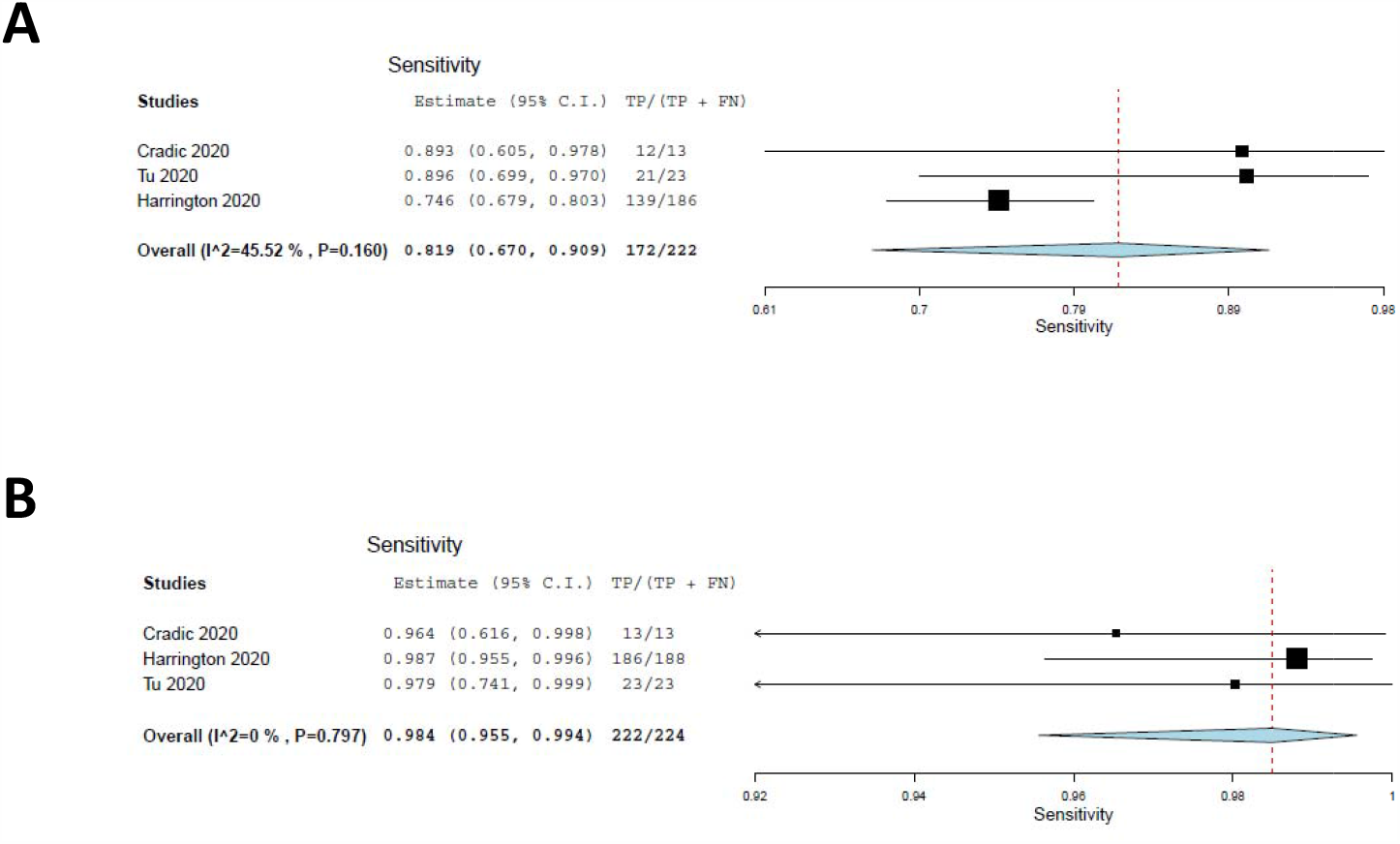
Forest plots demonstrating the three studies with low risk of patient selection bias utilized in the meta-analysis. Panel **A** demonstrates the sensitivity of ID NOW as compared with the reference standard, and the overall sensitivity was estimated to be 82% with a lower 95% confidence bound at 67% and an upper bound of 91%. Panel **B** demonstrates the sensitivity of RT-PCR and is estimated to be 98% with a 95% CI of 96-99%.

## Discussion

We conducted a large clinical evaluation of the ID NOW isothermal PCR system in a low-prevalence population and found that the ID NOW system had a positive percent agreement of 91% and a negative percent agreement of 100% compared to the Hologic Panther RT-PCR system. Several features which distinguish this study from those included in the systematic review are worth noting. The first is that the time from specimen collection to ID NOW testing was 15 minutes or less for most individuals tested. None of the studies meeting criteria for inclusion in the systematic review had such a short collection-to-testing time. A second feature of the current clinical evaluation, shared by only two of the studies included in our systematic review, was that the sample was based on a subject group that resulted from an attempt to enroll virtually every patient who walked through the door.

Most of the variation in performance reported for the ID NOW system seems to result from the differences in recruitment strategies employed in these studies. Peak viral loads and transmission risk for SARS-CoV-2 are found in symptomatic patients at symptom onset and then fall throughout the course of disease. Because RT-PCR assays have a limit of detection that are several orders of magnitude lower than that of the isothermal PCR ID NOW assay, one would expect them to remain positive for significantly longer times after the time of peak viral load. The use of “convenience samples,” particularly populations including patients who have been hospitalized after a diagnosis of COVID-19, may include more patients who are past their period of peak viral load compared to a sample of ambulatory patients first presenting for evaluation—such as those in our study who appeared for testing because of recent symptom onset. The two studies that met inclusion criteria for our review which had the lowest positive percent agreement between ID NOW and RT-PCR both included hospitalized patients,^13,17^ although another study with very low concordance did not.^14^

The conclusions from our clinical study are limited by a relatively small number of positive cases; nonetheless the high level of agreement with RT-PCR suggests that ID NOW is highly effective at identifying, or excluding, SARS-CoV-2 in a symptomatic ambulatory patient population. The systematic review and meta-analysis generally support this conclusion, although they suggest a somewhat lower positive percent agreement with RT-PCR.

Under the conditions of the current clinical study (population prevalence of 2.36%), the positive and negative predictive value of the ID NOW test were 100% and 99.8% (99.2% to 99.9%), respectively. At a prevalence of 10% in the tested population, the positive and negative predictive value are 100% and 99% (96.49% to 99.74%) respectively. Using the 82% estimate from our meta-analysis in a 2% positive population yields a negative predictive value of 99.6% (99.4 to 99.7%), which drops to 98.0% (97.1 to 98.7%) in a population with 10% disease prevalence. At the lower 95% confidence limit of the meta-analysis (67%), negative predictive value remains acceptable at 99.2% (99-99.4%) for a population prevalence of 2.3%. It becomes more marginal at 96.5% (95.4%-97.3%) when the prevalence of disease in the tested population goes to 10% or higher.

The data from our clinical study does not provide information on the potential utility of ID NOW in testing an asymptomatic patient population, since no positive cases were identified among the enrolled asymptomatic patients. Comparison of RT-PCR cycle numbers between symptomatic and asymptomatic ambulatory outpatients from The Everett Clinic suggests that the viral load for symptomatic patients is generally higher than for asymptomatic patients (Figure 3). This observation, which has also been reported elsewhere^18^, raises the possibility that ID NOW may miss infections in the asymptomatic infected population. On the other hand, the observation that specimens that demonstrate high Ct values are unlikely to be successfully cultured raises the possibility that many of these patients are less likely to transmit the infection, although the relationship between the ability to culture virus and infectivity has yet to be demonstrated for SARS-CoV-2.

**Figure 3:**
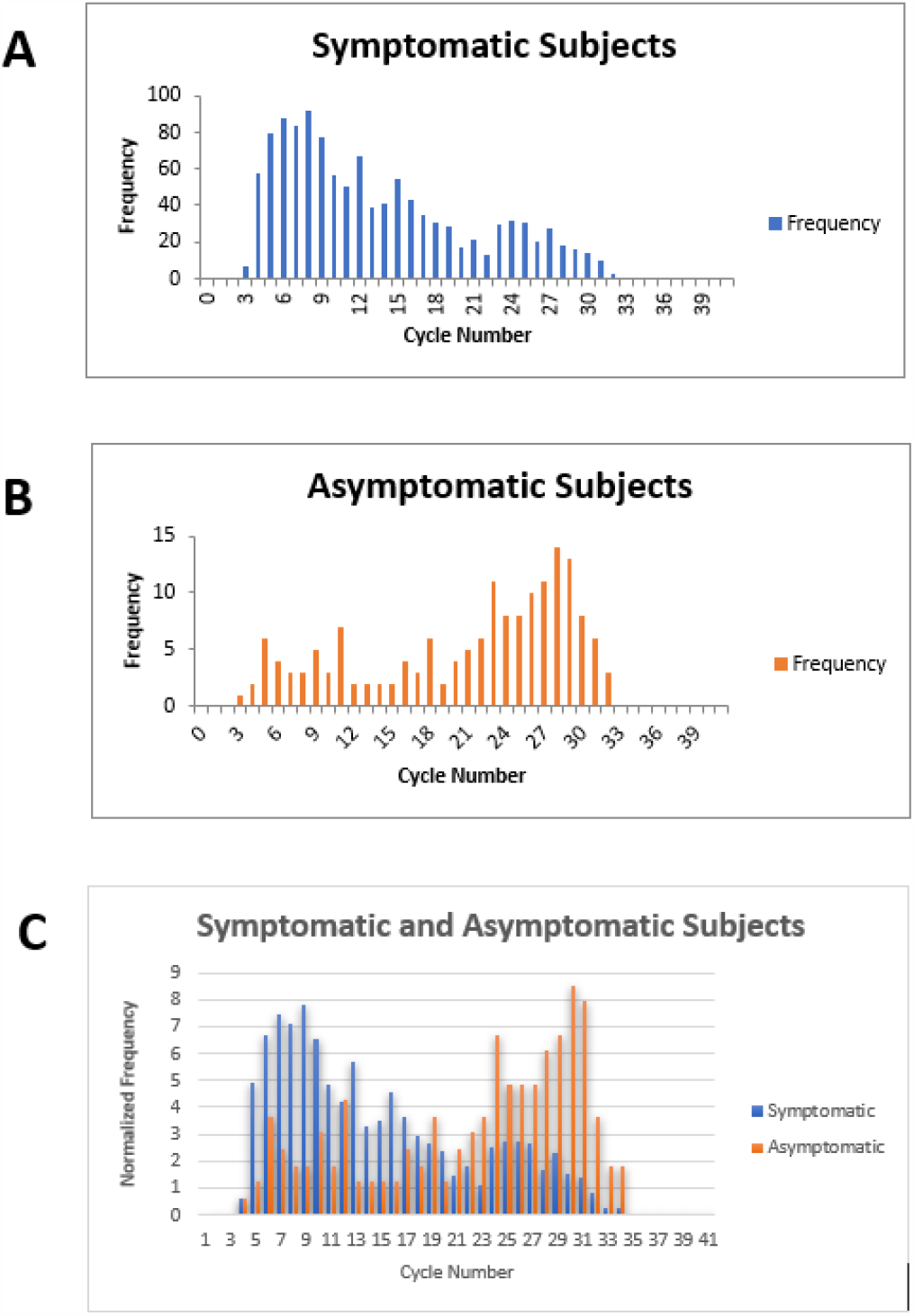
Cycle number distributions for 1182 symptomatic (panel **A**) and 164 asymptomatic (panel **B**) patients who tested positive for SARS-CoV-2 between July 14^th^ through November 16^th^, 2020, using the Abbott m2000 assay at The Everett Clinic. For patients with multiple tests, only the first positive test is included. In panel **C**, data for each group of patients has been normalized so that the sum of all bins is 100, allowing better comparison of the distributions. The Abbott m2000 cycle number is generally about 10 cycles less than the Ct reported for PCR assays on other devices.

Point of care testing has substantial advantages over laboratory-based testing when a patient presents with symptoms characteristic of COVID-19. Patients who are SARS-CoV-2 positive can be asked to isolate immediately, and patients who test negative can be reassured or retested using a more sensitive test, depending on clinical judgement. Although the performance of ID NOW in an asymptomatic population has not been established, and caution may be appropriate when using ID NOW with a high-risk population, increased frequency of testing, together with a rapid turnaround time, are likely to have greater impact on population health outcomes than are differences in test sensitivity^5,19^. In addition, the ID NOW system provides excellent negative predictive value in symptomatic ambulatory patients, particularly when the population prevalence of SARS-CoV2 is low. It thus provides a speedy and effective alternative to laboratory-based RT-PCR methods under many clinical circumstances.

## Data Availability

Data included in manuscript.

## Acknowledgments

The authors wish to acknowledge the contributions of Garrett Galbreath, Lorraine Bell, Susan Spanos, Anne Hartman, Amanda Wells, Kim Gangloff, Jeremy Norris, and Korie Packwood (from The Everett Clinic laboratory, analytical and electronic medical records staff), The Everett Clinic nurses, medical assistants, staff, providers and patients who graciously contributed to this study.

## Supplementary Materials

### Brief descriptions and confusion matrixes for each study

Computations using multiple samples to compute the composite reference standard are not shown.

#### Lephart^13^

This paper reports a comparison of Abbott ID NOW, Abbott m2000, Diasorin Simplexa and Cepheid Xpert Xpress using 75 nasopharyngeal (NP) and anterior nares (AN) swabs obtained from patients presenting in the ED and 13 from recovering COVID-positive inpatients. NP swabs were transported to a central laboratory and tested with the Simplexa, following which residual specimens were tested within 24 hours on the m2000 and Xpert Xpress devices. Nasal swabs were collected in parallel, transported dry to the laboratory, stored at 4°C and tested by ID NOW within 24 hours. The potential for patient selection bias is unclear because of the inclusion of recovering hospitalized subjects. Storage at 4°C for up to 24 hours is not consistent with current Abbott ID NOW instructions for use (IFU), resulting in unclear, though possibly insignificant risks for index test and flow and timing biases.

Data in the confusion matrix below, and in Table 2, reflects comparison of ID NOW with the Cepheid Xpert Xpress assay. Investigators noted that positive agreement was higher in patients with low m2000 cycle numbers.

Investigators also determined the limits of detection (LOD) for ID NOW and the m2000 assay, finding a LOD of 262 copies/mL for ID NOW, and 32.5 copies/mL for the m2000.

|  |  | **ID NOW** |  |
| --- | --- | --- | --- |
|  |  | Positive | Negative |
| **Cepheid XpertXpress** | Positive | 12 | 15 |
|  | Negative | 0 | 60 |

#### Basu^14^

The investigators obtained 101 paired foam AN (both nares) and NP swabs from patients presenting to the emergency department of a New York City Hospital between 22 and 24 April 2020. All swabs were transported to the laboratory at room temperature, while NP swabs were transported in VTM. The dry nasal swabs were tested within 2 hours of collection or kept at 4 to 8°C for up to 24 hours before testing on the ID NOW platform. NP swabs were tested on the Cepheid Xpert Xpress. Since no information is given about recruitment strategy, the risk of patient selection bias is unclear. Storage at 4-8°C for up to 24 hours is not consistent with current Abbott ID NOW instructions for use, resulting in unclear, though possibly insignificant risks for index test and flow and timing biases.

Investigators noted that all 6 patients with Xpert Xpress N2 Ct value of 33.5 tested positive by ID Now, while the positive percent agreement dropped substantially for cases in which the Xpert Xpress Ct value was higher.

|  |  | ID NOW |  |
| --- | --- | --- | --- |
|  |  | Positive | Negative |
| Cepheid Xpert Xpress | Positive | 17 | 14 |
|  | Negative | 1 | 69 |

#### Cradic^15^

NP, oropharyngeal (OP) and AN swabs were obtained prospectively from 182 consenting patients seen in an emergency department. NP swabs were placed in viral transport media (VTM) and transported to the lab for testing both by Diasorin Simplexa and Abbott ID NOW, With AN and OP swabs were tested by ID NOW. Risk of patient selection bias is low, but there is a lack of information regarding specimen flow and timing; thus, the risk of index test bias, and flow and timing bias is unclear.

Investigators used serial dilution (in VTM) of patient specimens to assess relative sensitivity of ID NOW compared with Roche Cobas and Diasorin Simplexa; the results suggest that ID NOW has a limit of detection about ten-fold higher than that of the Diasorin assay, and 100-fold higher than that of the Roche assay.

|  |  | ID NOW |  |
| --- | --- | --- | --- |
|  |  | Positive | Negative |
| Diasorin Simplexa | Positive | 12 | 1 |
|  | Negative | 0 | 169 |

#### Harrington^20^

Paired foam nasal swabs (NS) and NP swabs were obtained from 524 symptomatic subjects presenting consecutively at three emergency departments (ED) and two immediate care centers. Both nasal swabs (dry) were tested locally using ID NOW, and NPS were transported to a central laboratory and NPS (in VTM) were transported to a central laboratory and tested using the Abbott m2000. The risk of patient selection bias was rated to be low; since no information was given regarding the interval between specimen acquisition and ID NOW testing, the risk of index test bias and flow and timing bias were rated unclear.

Harrington estimated the LOD for the ID NOW assay to be 3225 copies/mL, based on the package insert which reports an LOD of 100 genome-equivalents/ml.

|  |  | ID NOW |  |
| --- | --- | --- | --- |
|  |  | Positive | Negative |
| Abbott m2000 | Positive | 139 | 47 |
|  | Negative | 2 | 336 |

#### Jin^21^

As part of a larger study, investigators compared the results from 52 dry swabs tested with the ID NOW system with those of paired NP specimens tested on the Roche COBAS system. Details regarding patient recruitment and the patient to test time for were not provided. For this reason, the risk of patient recruitment bias, index test bias and flow and timing bias are all considered to be unclear.

Investigators also estimated that the LOD for the ID NOW assay is 16-fold higher than that of the Roche Cobas assay.

|  |  | ID NOW |  |
| --- | --- | --- | --- |
|  |  | Positive | Negative |
| Roche COBAS | Positive | 4 | 2 |
|  | Negative | 0 | 46 |

#### Mitchell^16^

Previously tested residual positive and negative nasopharyngeal patient samples collected in VTM and stored at −80C were tested using the ID NOW EUA assay. Risk of recruitment bias is high because the specimens were selected by investigators, in part upon the basis of previous RT-PCR testing results. RT-PCR was performed using either a CDC or a New York State assay which had been granted Emergency Use authorization by the FDA. The risk of index test bias is high because dry swabs were not employed, and the risk of flow and timing bias is unclear.

Investigators correlated the ID NOW performance with the Ct obtained during PCR. Although a 72% false negative rate was found for patients whose Ct ranged from 35 to 40, no false negatives were identified for patients with a Ct below 35.

|  |  | ID NOW |  |
| --- | --- | --- | --- |
|  |  | Positive | Negative |
| RTPCR – either CDC or New York EUA | Positive | 33 | 13 |
|  | Negative | 0 | 15 |

#### Moore^22^

NP swabs in VTM were collected from a mix of ambulatory and hospitalized patients, some who were in the ICU. Some patient specimens were obtained consecutively, while, additional specimens were obtained by including samples in which SARS-CoV-2 RNA was detected by RT-PCR, and others were included in which virus had not been detected. Risk of patient selection bias is high, while the risks of index test and flow and timing biases are unclear. Specimens were analyzed within 72 hours of collection and were held refrigerated at 4°C if all testing could not be completed on the same day.

In a separate evaluation, NP swabs were collected from 97 symptomatic emergency department patients who had negative results from a dry nasal swab tested at the point of care by ID NOW. These NP swabs were subsequently evaluated by R-TPCR on the Abbott m2000 system. SARS-CoV-2 RNA was detected in 13 of these NP swabs, or 13.4% of the total. The median Ct for these ID NOW – negative / m2000 - positive cases was 19.82.

|  |  | ID NOW |  |
| --- | --- | --- | --- |
|  |  | Positive | Negative |
| Abbott m2000 | Positive | 94 | 31 |
|  | Negative | 0 | 73 |

#### Rhoades^23^

A convenience sample of ninety-six clinical remnant specimens (11 supervised self-collected nasal swabs in normal saline and 85 provider-collected NP swabs in VTM) that had previously tested positive for SARS-CoV2 by RT-PCR were retested using ID Now, Diasorin Simplexa and a modified CDC LDT method. The study was rated as having a high risk of patient selection bias, and unclear risk of both index test bias and flow and timing bias.

|  |  | ID NOW |  |
| --- | --- | --- | --- |
|  |  | Positive | Negative |
| Diasorin Simplexa | Positive | 90 | 6 |
|  | Negative | 0 | 0 |

#### Smithgall^3^

Investigators compared performance of Roche Cobas 6800, Cepheid Xpert and Abbott ID NOW assays using 88 residual NP swabs previously confirmed as positive that were chosen to represent the full range of observed Ct values, and 25 NP swabs previously confirmed as negative, all of which had been held at 4°C in VTM for no more than 48 hours prior to testing. Risk of patient selection bias is rated as high due to the modified case-control design, and risk of index test bias and flow and timing bias are rated as unclear. The table below shows the confusion matrix for ID NOW compared to the Roche COBAS 6800, but the composite values shown in Table 2 of the paper reflect the fact that the Cepheid Xpert identified two cases not identified by COBAS, while COBAS identified one case not identified by Xpert.

The authors demonstrated that agreement between ID NOW and RTPCR was perfect for Cobas Ct≤30, but fell off dramatically at higher Ct.

|  |  | ID NOW |  |
| --- | --- | --- | --- |
|  |  | Positive | Negative |
| Roche COBAS | Positive | 65 | 23 |
|  | Negative | 0 | 25 |

#### Thwe^17^

Investigators compared results from 182 paired dry NP swabs on ID NOW, and NP swabs in VTM, from symptomatic hospitalized and emergency department patients. Dry NP swabs were transported to the ID NOW testing area within 2 hours of collection, but total time to analysis was not reported. We rate the risk of patient recruitment bias to be high due to the inclusion of hospitalized inpatients. We rate the risk of flow and timing bias, and index test bias, as unclear. PCR was carried out using any of several systems, including The real-time Abbott RealTime SARS-CoV-2 (Abbott Park, IL, USA), Panther Fusion® SARS-COV-2 (San Diego, CA, USA), and Cepheid Xpert® Xpress SARS-CoV-2 (Sunnyvale, CA, USA) and a laboratory-developed test (LDT).

|  |  | ID NOW |  |
| --- | --- | --- | --- |
|  |  | Positive | Negative |
| RT-PCR | Positive | 8 | 7 |
|  | Negative | 0 | 167 |

#### Zhen^24^

This study included 108 NP swabs from symptomatic patients, 20 of which were collected prospectively and 88 of which were taken from a collection of frozen specimens (−80C) that had been previously tested. All samples were tested using the Hologic Panther Fusion SARS-CoV-2 assay, the Abbott ID NOW assay and the Cepheid Xpert Xpress assay. The prospective 20 specimens were processed fresh on each platform at the time of patient testing. Risk of patient selectin bias was rated as unclear. Risk of index test bias was considered to be unclear, due to use of frozen specimens, and risk of flow and timing bias was also rated as unclear.

All false-negative ID NOW results were associated with Hologic Panther Ct ≥ 32.

Authors also performed a limit-of-detection analysis and found an LOD for ID NOW of 20,000 copies/ml, with LOD for Xpert Xpress of 100 copies/ml, and 1000 copies/ml for GenMark ePlex.

|  |  | ID NOW |  |
| --- | --- | --- | --- |
|  |  | Positive | Negative |
| Hologic Panther Fusion | Positive | 50 | 7 |
|  | Negative | 0 | 50 |

#### Comer^25^

This study sampled all COVID-19 symptomatic prospective hospital admissions with combined nasopharyngeal (NP) and oropharyngeal (OP) swabs in the ED with the Abbott ID NOW and tested them immediately in the emergency department, if negative, recollect expeditiously and test on a Becton Dickinson BD MAX. Retesting occurred within a few hours. The risk of patient selection bias appears to be high, based upon considering a group that is being considered for hospitalization, rather than the general population of symptomatic patients possibly suffering from SARS-CoV-2 infection. The risks of index test bias and flow and timing bias appear to be low.

|  |  | ID NOW |  |
| --- | --- | --- | --- |
|  |  | Positive | Negative |
| **Becton Dickinson BD MAX** | Positive | 0 | 1 |
|  | Negative | 0 | 116 |

#### Ghofrani^26^

Investigators employed a “convenience sample” that included patients who had a RT-PCR sample collected close to the time of presentation followed by a re-swab for ID NOW, and those who were already known to be PCR-positive and whose residual NP samples were tested by IDNOW. Some specimens employed in the ID NOW testing were dry, while others were transported in UTM. RT-PCR testing was conducted at one of several different laboratories, and the specific tests utilized were not reported. Risk of both patient selection bias and of flow and timing bias considered to be high. The risk of index test bias is rated as unclear.

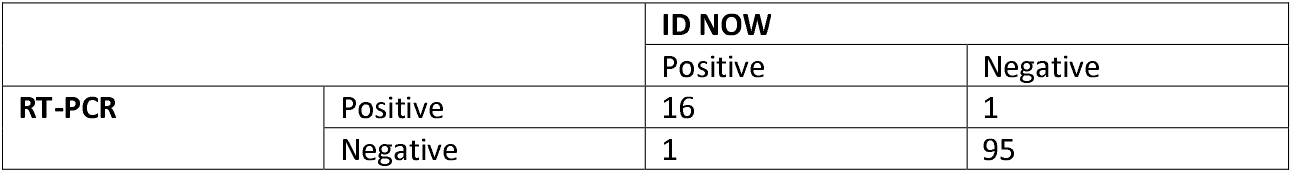

|  |  | ID NOW |  |
| --- | --- | --- | --- |
|  |  | Positive | Negative |
| **RT-PCR** | Positive | 16 | 1 |
|  | Negative | 1 | 95 |

#### SoRelle^27^

This letter reports a study in which ID NOW as compared with Cepheid Xpert Xpress on 59 saliva samples from symptomatic subjects. Details regarding collection environment and saliva transport are not provided. Investigators also compared saliva tested using the ID NOW system with NPS testing using RT-PCR (Abbott m2000); this data is not included in the present review. Results from a single test reported as “invalid” on the ID NOW platform are not included in the confusion matrix below.

There is an uncertain risk of patient recruitment bias due to the lack of information. We have rated the risk of index test bias and flow and timing bias as low, based on the author’s assertion that all testing was performed in accordance with the manufacturer’s instructions.

Investigators noted that most ID NOW false negative results occurred in patients tested ≥2 weeks after symptom onset. They estimated the LOD for the ID NOW assay at 2000 copies/mL.

|  |  | **ID NOW** |  |
| --- | --- | --- | --- |
|  |  | Positive | Negative |
| **Cepheid Xpert Xpress** | Positive | 23 | 0 |
|  | Negative | 0 | 35 |

**Supplemental Figure 1:**
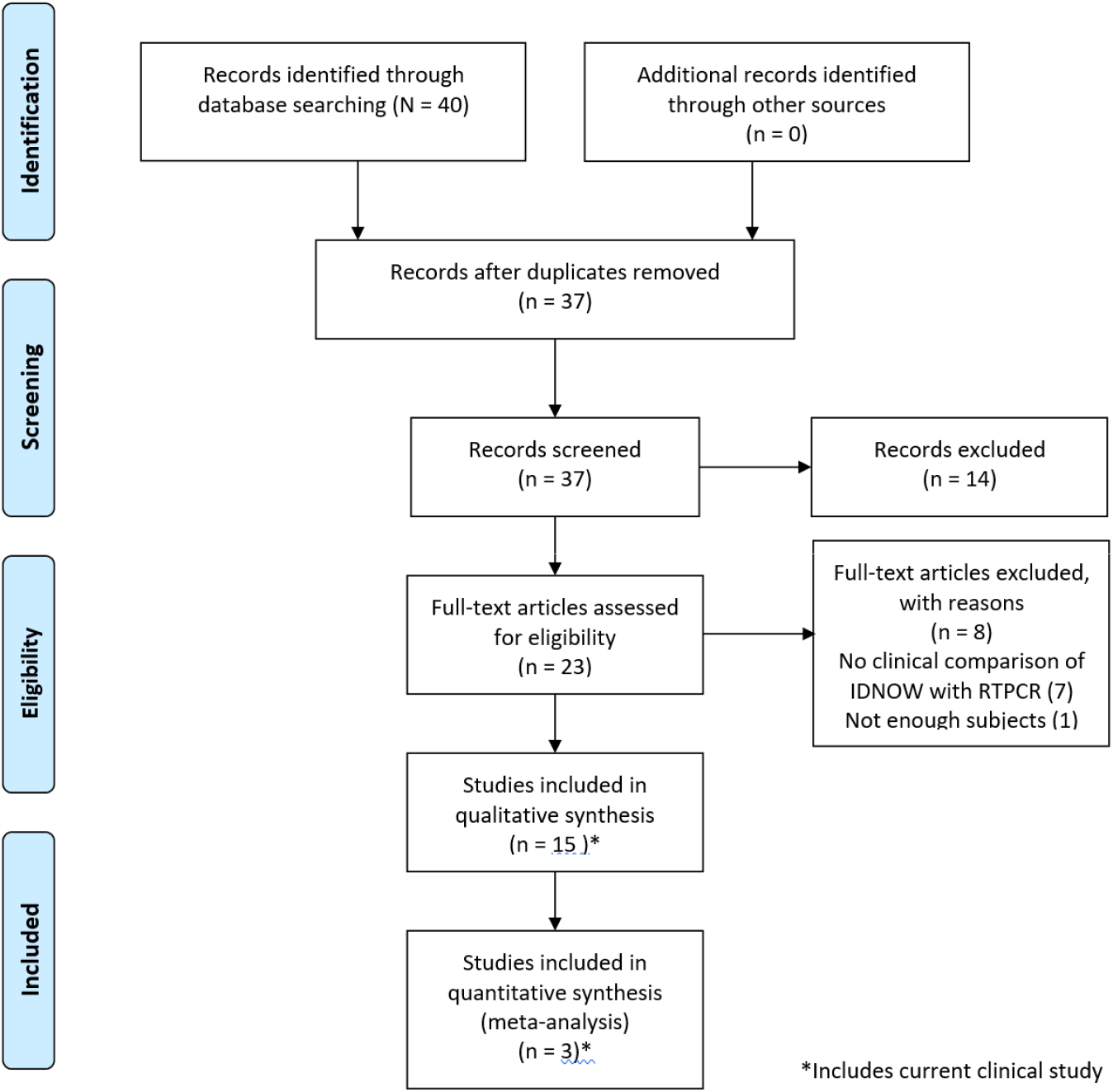
PRISMA 2009 Flow Diagram detailing the studies that were identified, screened, deemed eligible and finally included in the analysis. Note that the data from the current clinical evaluation has been included in the analysis.

